# Modifications to student quarantine policies in K–12 schools implementing multiple COVID-19 prevention strategies restores in-person education without increasing SARS-CoV-2 transmission risk, January-March 2021

**DOI:** 10.1101/2022.03.18.22272631

**Authors:** Patrick Dawson, Mary Claire Worrell, Sara Malone, Stephanie A. Fritz, Heather P. McLaughlin, Brock K. Montgomery, Mary Boyle, Ashley Gomel, Samantha Hayes, Brett Maricque, Albert M. Lai, Julie A. Neidich, Sarah C. Tinker, Justin S. Lee, Suxiang Tong, COVID-19 Response Fieldwork and Laboratory Teams, Rachel C. Orscheln, Rachel Charney, Terri Rebmann, Missouri School District Data and Coordination Group, Jon Mooney, Catherine Rains, Nancy Yoon, Machelle Petit, Katie Towns, Clay Goddard, Spring Schmidt, Lisa C. Barrios, John C. Neatherlin, Johanna S. Salzer, Jason G. Newland

## Abstract

**Objective:** To determine whether modified K–12 student quarantine policies that allow some students to continue in-person education during their quarantine period increase schoolwide SARS-CoV-2 transmission risk following the increase in cases in winter 2020-2021.

**Methods:** We conducted a prospective cohort study of COVID-19 cases and exposures among students and staff (n=65,621) in 103 Missouri public schools. Participants were offered free, saliva-based RT-PCR testing. An adjusted Cox regression model compared hazard rates of school-based SARS-CoV-2 infections between schools with a modified versus standard quarantine policy.

**Results:** From January–March 2021, a projected 23 (1%) school-based transmission events occurred among 1,636 school close contacts. There was no difference in the adjusted hazard rates of school-based SARS-CoV-2 infections between schools with a modified versus standard quarantine policy (hazard ratio=1.00; 95% confidence interval: 0.97–1.03).

**Discussion:** School-based SARS-CoV-2 transmission was rare in 103 K–12 schools implementing multiple COVID-19 prevention strategies. Modified student quarantine policies were not associated with increased school incidence of COVID-19. Modifications to student quarantine policies may be a useful strategy for K–12 schools to safely reduce disruptions to in-person education during times of increased COVID-19 community incidence.

## Introduction

The coronavirus disease 2019 (COVID-19) pandemic substantially impacted the operation of kindergarten through grade 12 (K–12) schools in the United States, with many schools switching from in-person instruction to part- or full-time virtual learning during the 2020–2021 school year (1). Virtual instruction can be an effective learning model for some students, but unequal access to computers, internet services, and childcare services may put some children at an educational disadvantage (2). Disruptions to in-person instruction may also affect students beyond educational attainment, as K–12 schools often provide critical services (e.g., nutritional, physical, and mental health support) to students and their families (3). Therefore, schools nationwide have sought strategies to safely operate and maintain in-person learning during the COVID-19 pandemic.

The U.S. Food and Drug Administration (FDA) has authorized the Comirnaty/ Pfizer-BioNTech COVID-19 Vaccine for use in children ages 5 years and older through an Emergency Use authorization (ages 5-15 years) and a full authorization for ages 16 and above (4). As of March 14, 2022, 34% of the population 5-11 years and 70% of the population 12-17 years had received at least one dose of a COVID-19 vaccine (5). Vaccine hesitancy and inequitable access to vaccine mean not all communities have high coverage of COVID-19 vaccination (6, 7). Thus, layered prevention strategies continue to be essential to prevent school-based SARS-CoV-2 transmission for K–12 school students when there are increased COVID-19 Community Levels (8): implementing a universal masking policy, ensuring physical distancing in classrooms, increasing classroom ventilation with outdoor air, screening testing, having a robust case identification and contact tracing system, and following local isolation and quarantine guidance (9). Under a standard quarantine policy, students typically must forfeit all in-person activities including in-person instruction for 7–14 days after their last exposure (10).

However, some schools have implemented modifications to standard quarantine guidance to minimize disruptions to in-person learning for students if the exposure is deemed low risk. In Greene County, Missouri, K–12 schools implemented a modified quarantine policy permitting student close contacts of a person having COVID-19 to attend school in person during their quarantine period if: they were aged ≤18 years, their only exposure was in a classroom, they did not have direct physical contact for ≥15 minutes in one 24-hour period with the person having COVID-19, and both the student close contact and the person having COVID-19 had worn masks appropriately during the exposure event (11). In a two-week investigation in Greene County in December 2020, an estimated 240 days of in-person instruction were saved for 30 students participating in the modified quarantine (12). Additionally, none of the students participating in the modified quarantine were identified as having a SARS-CoV-2 infection during the 14 days following exposure through testing or symptom monitoring (12).

Layered COVID-19 prevention strategies in K–12 schools can limit school-based SARS-CoV-2 transmission despite concurrent high community incidence (9, 12-15), unless there are lapses in implementation or adherence (16, 17). However, the effect of individual prevention strategies on school-based transmission has not been thoroughly assessed in previous reports. Furthermore, less is known about the impact that modifications to quarantine policies have on school-based transmission risk and schoolwide COVID-19 incidence.

To better understand the effects of COVID-19 prevention strategies and modifications to quarantine policies on school-based SARS-CoV-2 transmission, we conducted a two-month investigation in Missouri public schools. The objectives were to 1) measure the frequency of school-based SARS-CoV-2 transmission; 2) quantify the relative risks of school-based transmission among schoolwide COVID-19 policies; and 3) compare the schoolwide incidence between schools implementing a modified quarantine and schools following standard quarantine.

## Methods

During January 25–March 21, 2021, we worked with school officials in 103 schools across six Missouri public school districts in Greene County (57 schools; districts A–C) and St. Louis County (46 schools; districts D–F), with an estimated 65,621 students and staff (S1 Table). During the investigation, districts A–C schools had implemented a modified quarantine policy while districts D–F schools followed standard quarantine (11, 18, 19). The project was reviewed and approved by the Washington University in St. Louis Institutional Review Board and conducted consistent with applicable federal law and U.S. Centers for Disease Control and Prevention (CDC) policy (45 C.F.R. part 46, 21 C.F.R. part 56; 42 U.S.C. Sect. 241(d); 5 U.S.C. Sect. 552a; 44 U.S.C. Sect. 3501 et seq.).

School officials were notified of students and staff members who received a positive SARS-CoV-2 nucleic acid amplification test (NAAT) or antigen test, typically ≤2 days after the laboratory result. For persons having COVID-19 who had been physically present in school, at a school-associated event (e.g., school athletics, extracurricular activities), or on a school bus while potentially infectious (starting two days before symptom onset or collection of their first positive test specimen), school officials conducted contact tracing to identify their school-based close contacts. A close contact was defined as someone who was ≤6 feet away from a person with COVID-19 for ≥15 minutes in one 24-hour period. In districts A–C, school officials determined if student close contacts (hereafter, contacts) met criteria for a modified quarantine. School officials followed everyone through completion of their isolation or quarantine period, including whether contacts received a positive NAAT or antigen test.

After the school case notification and contact tracing is completed, we asked eligible individuals to participate in an enhanced investigation consisting of a telephone interview and free saliva-based testing. Persons with COVID-19 were eligible for the enhanced investigation if they had been physically present in school, at a school-associated event, or on a school bus while potentially infectious within 14 days of recruitment by the investigation team; contacts were eligible if their most recent school-based exposure was within 14 days of recruitment. Contacts were ineligible if they lived with the person with COVID-19 from the school-based exposure. Participants provided oral agreement to participate, and parents/guardians provided oral agreement for children aged <18 years.

For the enhanced investigation, a trained interviewer conducted a standardized telephone interview addressing clinical symptoms; school, community, and household exposures; and demographics. For children aged 12–17 years, a parent/guardian and/or the child was interviewed; for children aged <12 years, only a parent/guardian was interviewed. Upon completion of their quarantine period, participating contacts were reinterviewed about symptoms, additional exposures and activities, and SARS-CoV-2 testing. Saliva specimens were collected from persons with COVID-19 soon after recruitment and from contacts 5–14 days after their last school-based exposure. Specimens were tested for SARS-CoV-2 by real-time reverse transcription polymerase chain reaction (RT-PCR) as previously described.(12) Full genome sequences were generated from RT-PCR–positive saliva specimens at CDC (20).

For each school-based contact who received a positive test result from the enhanced investigation or elsewhere reported to school officials, we conducted a case determination process to assess the likelihood of the infection being from school-based transmission. Infections were classified as probable, possible, or unlikely school-based transmission using epidemiologic and sequencing data from case-contact pairs. School-based transmission was considered unlikely if the close contact lived in the same household as another person with COVID-19 ≤14 days before symptom onset or date of collection of their first positive specimen; their exposure, symptom, or testing timeline was not consistent with the known epidemiology of COVID-19; or the sequence generated from their specimen had >5 single nucleotide polymorphisms (SNPs) compared to the sequence generated from their school-based index case’s specimen. School-based transmission was considered possible if the close contact had non-household community exposure to a person with COVID-19 ≤14 days before symptom onset or date of collection of their first positive specimen and was considered probable if their only identified close contact was with the school-based person with COVID-19. If the sequence generated from the close contact’s specimen had ≤5 SNPs compared to the sequence generated from their school-based index case’s specimen, it was classified as probable school-based transmission. Classifications were made by at least two members of the investigation team. Discordant classifications were resolved by group discussion with at least one additional team member. In the absence of these data, classification defaulted to probable.

Data on school- and district-level student enrollment, staffing, demographics, and COVID-19 prevention measures were collected from school officials using a standardized survey. All contact tracing data from school officials were collected in Microsoft Excel and entered into a REDCap database (hosted by Washington University in St. Louis; version 9.5.5) along with data from the enhanced investigation. Data were cleaned and analyzed using R (R Core Team; version 3.6.1) and SAS (SAS Institute; version 9.4).

Descriptive analyses were conducted and two-sided tests for statistical significance were measured using Fisher’s exact test where appropriate. Relative risks of school-based SARS-CoV-2 transmission by schoolwide COVID-19 policies were computed using log-binomial regression and accounted for school-level cluster-correlated observations using generalized estimating equations with an independent correlation matrix. Adjusted models included a variable indicating whether the contact was a student or a staff member.

The percentage of asymptomatic contacts we tested who received a positive test result was extrapolated to contacts who were never tested to project the total number of cases (contacts who did not receive testing were presumed to be asymptomatic). Schoolwide COVID-19 crude incidence rates and Cox proportional hazard rates (using observed and projected total case numbers) were compared between schools with a modified versus standard quarantine policy. The approximate number of students and staff at each school whose attendance was 100% virtual or had COVID-19 ≤90 days before the start of the study were subtracted from the denominator. Those presumed to have been infected outside of school were censored at the day of their positive test specimen collection. Hazard ratios were adjusted for potential school-level confounding factors: quartiles of the percentage of students eligible to receive free or reduced-price lunch (as a proxy for school resources) and the school’s total number of persons with COVID-19 that attended school or a school-related event during the study period.

## Results

Of 103 participating schools, COVID-19 prevention strategy survey data were available for 100 (97%). S1 Fig describes the mitigation strategies being utilized in the schools. Virtual instruction was offered by 92% of schools, universal masking policies (face masks required for all students, teachers, staff, and visitors on school grounds) were implemented in 97% of schools, 94% of schools reported efforts to increase ventilation in classrooms, and desks were spaced at least three feet apart in all classrooms in 84% of schools.

From January 25 to March 21, 2021, a total of 1,864 students, teachers, and staff were identified by school officials through case identification and contact tracing, including 228 eligible persons with COVID-19 and 1,636 contacts (Table 1). Among contacts, 16 (1%) had two school-based exposures within the same 14-day window and 12 (1%) had a second school-based exposure after completion of their first quarantine period. Eligible cases and contacts were identified at 68 (66%) schools. The median number of contacts identified per school index person with COVID-19 was 6 (interquartile range, 2–11).

**Table 1.**
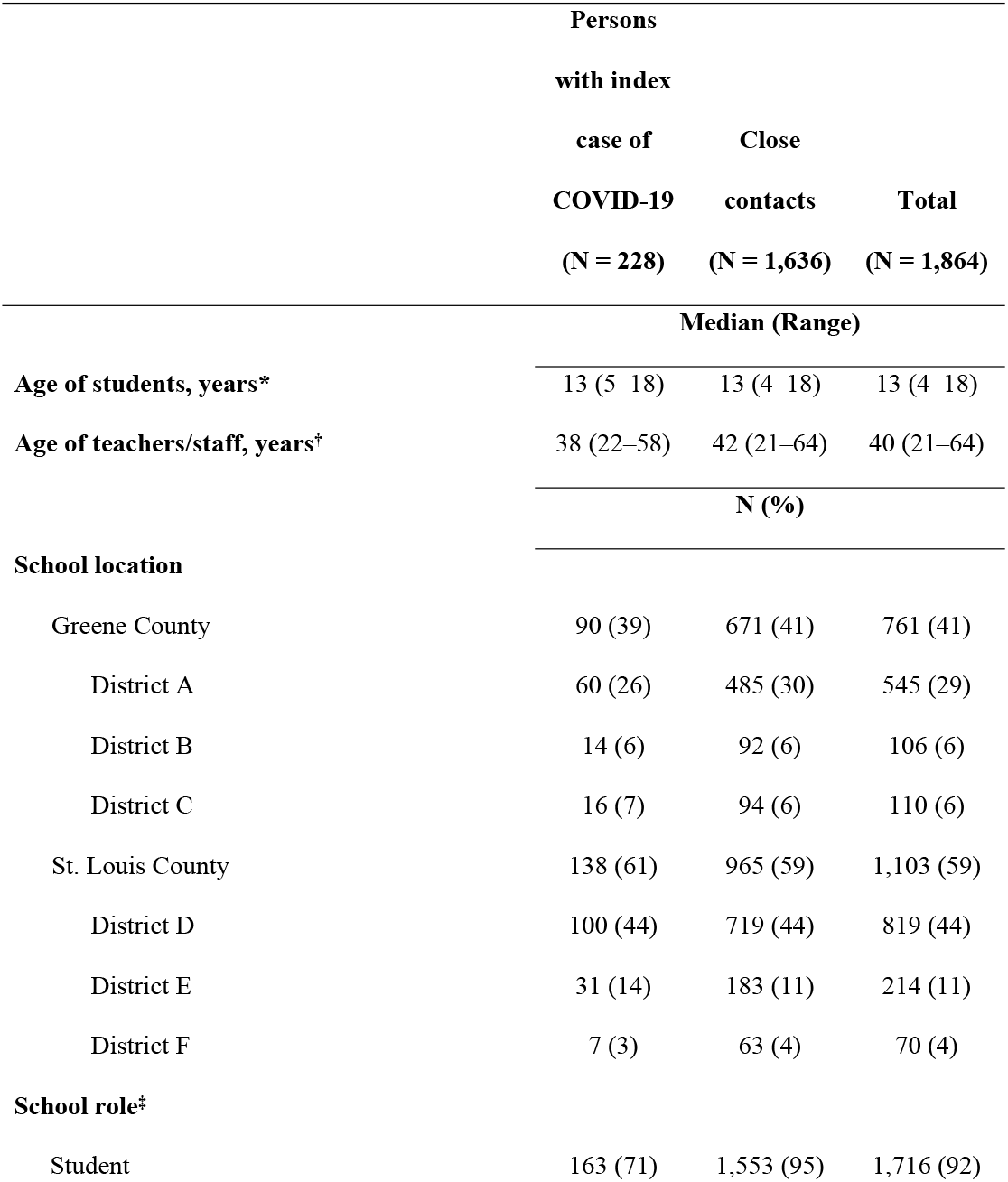
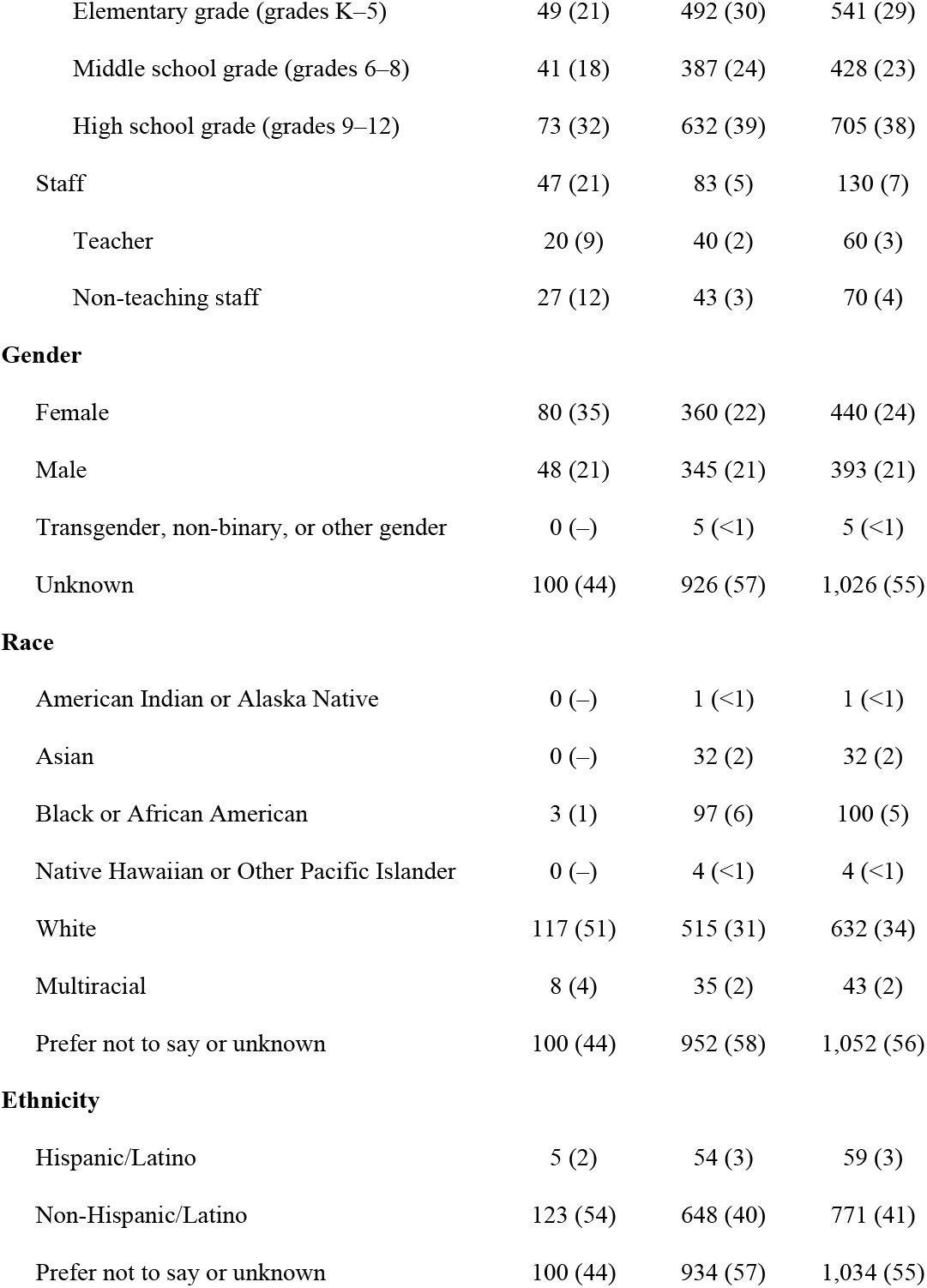
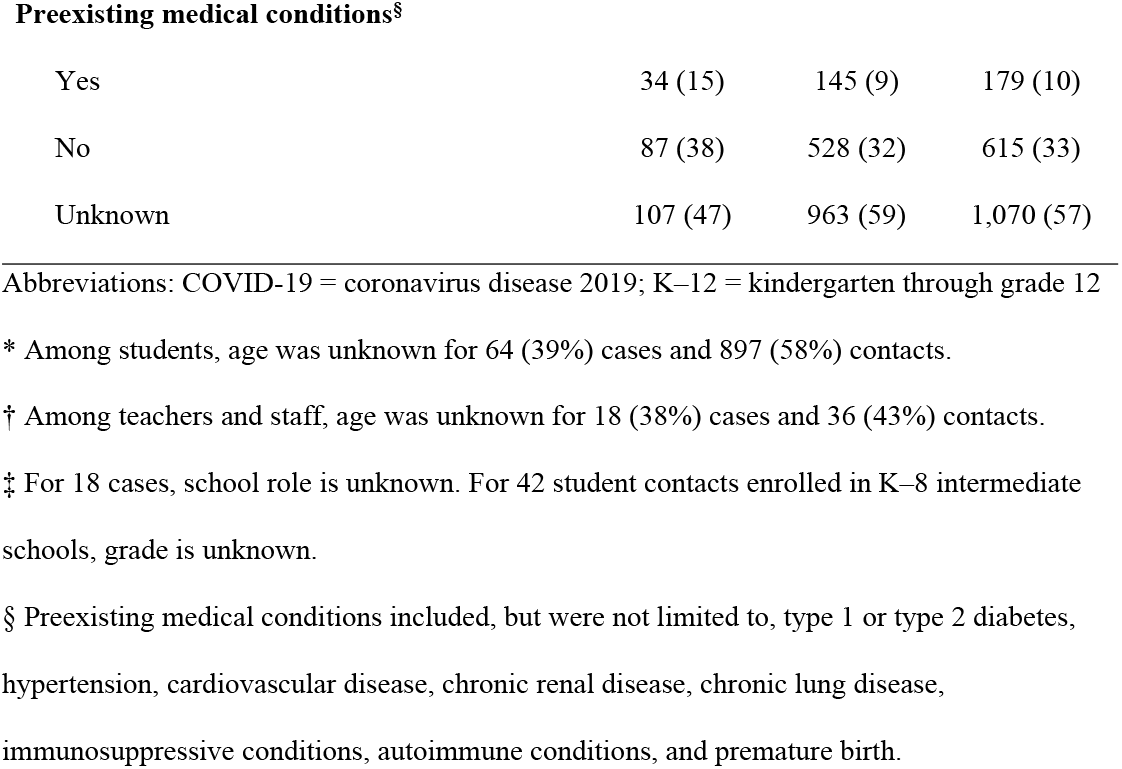
Characteristics of school-based persons with index cases of COVID-19 and close contacts from 103 K–12 schools, Greene and St. Louis Counties, Missouri, January 25– March 21, 2021.

|  | **Persons** |  |  |
| --- | --- | --- | --- |
|  | **with index** |  |  |
|  | **case of** | **Close** |  |
|  | **COVID-19** | **contacts** | **Total** |
|  | **(N = 228)** | **(N = 1,636)** | **(N = 1,864)** |
|  | **Median (Range)** |  |  |
| **Age of students, years\*** | 13 (5–18) | 13 (4–18) | 13 (4–18) |
| **Age of teachers/staff, years<sup>†</sup>** | 38 (22–58) | 42 (21–64) | 40 (21–64) |
|  | **N (%)** |  |  |
| **School location** |  |  |  |
| Greene County | 90 (39) | 671 (41) | 761 (41) |
| District A | 60 (26) | 485 (30) | 545 (29) |
| District B | 14 (6) | 92 (6) | 106 (6) |
| District C | 16 (7) | 94 (6) | 110 (6) |
| St. Louis County | 138 (61) | 965 (59) | 1,103 (59) |
| District D | 100 (44) | 719 (44) | 819 (44) |
| District E | 31 (14) | 183 (11) | 214 (11) |
| District F | 7 (3) | 63 (4) | 70 (4) |
| **School role<sup>‡</sup>** |  |  |  |
| Student | 163 (71) | 1,553 (95) | 1,716 (92) |
| Elementary grade (grades K–5) | 49 (21) | 492 (30) | 541 (29) |
| Middle school grade (grades 6–8) | 41 (18) | 387 (24) | 428 (23) |
| High school grade (grades 9–12) | 73 (32) | 632 (39) | 705 (38) |
| Staff | 47 (21) | 83 (5) | 130 (7) |
| Teacher | 20 (9) | 40 (2) | 60 (3) |
| Non-teaching staff | 27 (12) | 43 (3) | 70 (4) |
| **Gender** |  |  |  |
| Female | 80 (35) | 360 (22) | 440 (24) |
| Male | 48 (21) | 345 (21) | 393 (21) |
| Transgender, non-binary, or other gender | 0 (–) | 5 (<1) | 5 (<1) |
| Unknown | 100 (44) | 926 (57) | 1,026 (55) |
| **Race** |  |  |  |
| American Indian or Alaska Native | 0 (–) | 1 (<1) | 1 (<1) |
| Asian | 0 (–) | 32 (2) | 32 (2) |
| Black or African American | 3 (1) | 97 (6) | 100 (5) |
| Native Hawaiian or Other Pacific Islander | 0 (–) | 4 (<1) | 4 (<1) |
| White | 117 (51) | 515 (31) | 632 (34) |
| Multiracial | 8 (4) | 35 (2) | 43 (2) |
| Prefer not to say or unknown | 100 (44) | 952 (58) | 1,052 (56) |
| **Ethnicity** |  |  |  |
| Hispanic/Latino | 5 (2) | 54 (3) | 59 (3) |
| Non-Hispanic/Latino | 123 (54) | 648 (40) | 771 (41) |
| Prefer not to say or unknown | 100 (44) | 934 (57) | 1,034 (55) |
| **Preexisting medical conditions<sup>§</sup>** |  |  |  |
| Yes | 34 (15) | 145 (9) | 179 (10) |
| No | 87 (38) | 528 (32) | 615 (33) |
| Unknown | 107 (47) | 963 (59) | 1,070 (57) |

Abbreviations: COVID-19 = coronavirus disease 2019; K–12 = kindergarten through grade 12
\* Among students, age was unknown for 64 (39%) cases and 897 (58%) contacts.
† Among teachers and staff, age was unknown for 18 (38%) cases and 36 (43%) contacts.
‡ For 18 cases, school role is unknown. For 42 student contacts enrolled in K–8 intermediate schools, grade is unknown.
§ Preexisting medical conditions included, but were not limited to, type 1 or type 2 diabetes, hypertension, cardiovascular disease, chronic renal disease, chronic lung disease, immunosuppressive conditions, autoimmune conditions, and premature birth.

Of 1,636 contacts, 496 (30%) were tested for SARS-CoV-2, and 14 (3% of tested; 1% overall) received a positive test result. Among tested contacts, we tested 372 (75%) and 124 (25%) were tested elsewhere. Case determinations for the 14 contacts with a positive test result classified 11 as resulting from probable school-based transmission, one as possible, and two as unlikely. For subsequent analyses, probable and possible were combined for a total of 12 school-based transmission events. Six contacts who received a positive test result had paired whole genome sequencing data with their school-based index case, which confirmed epidemiologic linkages for four (67%; all identical or nearly identical sequences) and ruled out epidemiologic linkages for two (33%; classified as unlikely). One pair’s sequences, both assigned to the B.1.2 lineage, differed by 15 single nucleotide polymorphisms (SNPs). The other pair’s sequences, assigned to lineages B.1.2 and B.1.1.416, differed by 35 SNPs. The 12 school-based transmission events were generated by 11 school index cases (5%) in 10 different schools; a cluster in one school involved a transmission chain with one primary, one secondary, and two tertiary cases (Table 2).

**Table 2.**
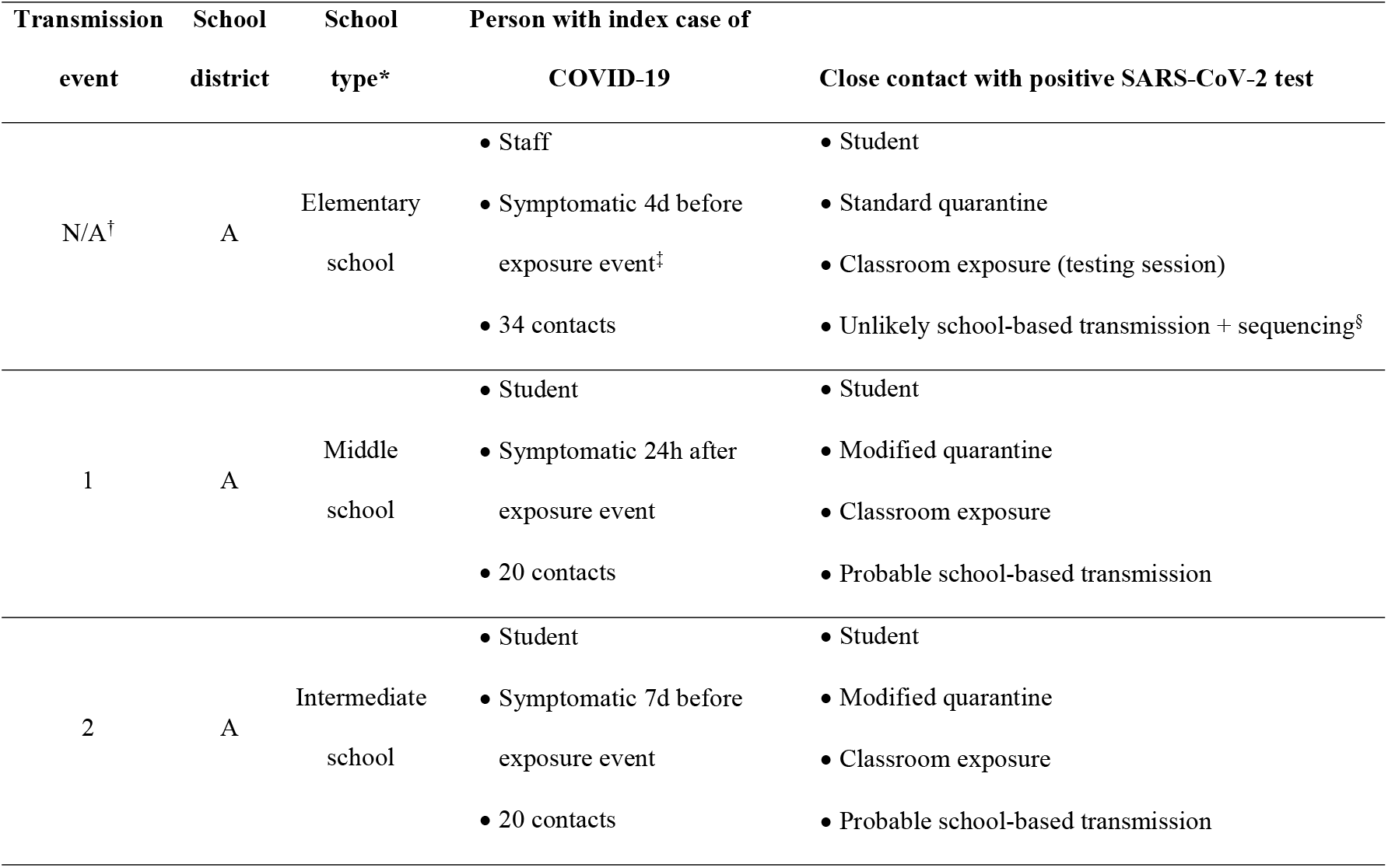
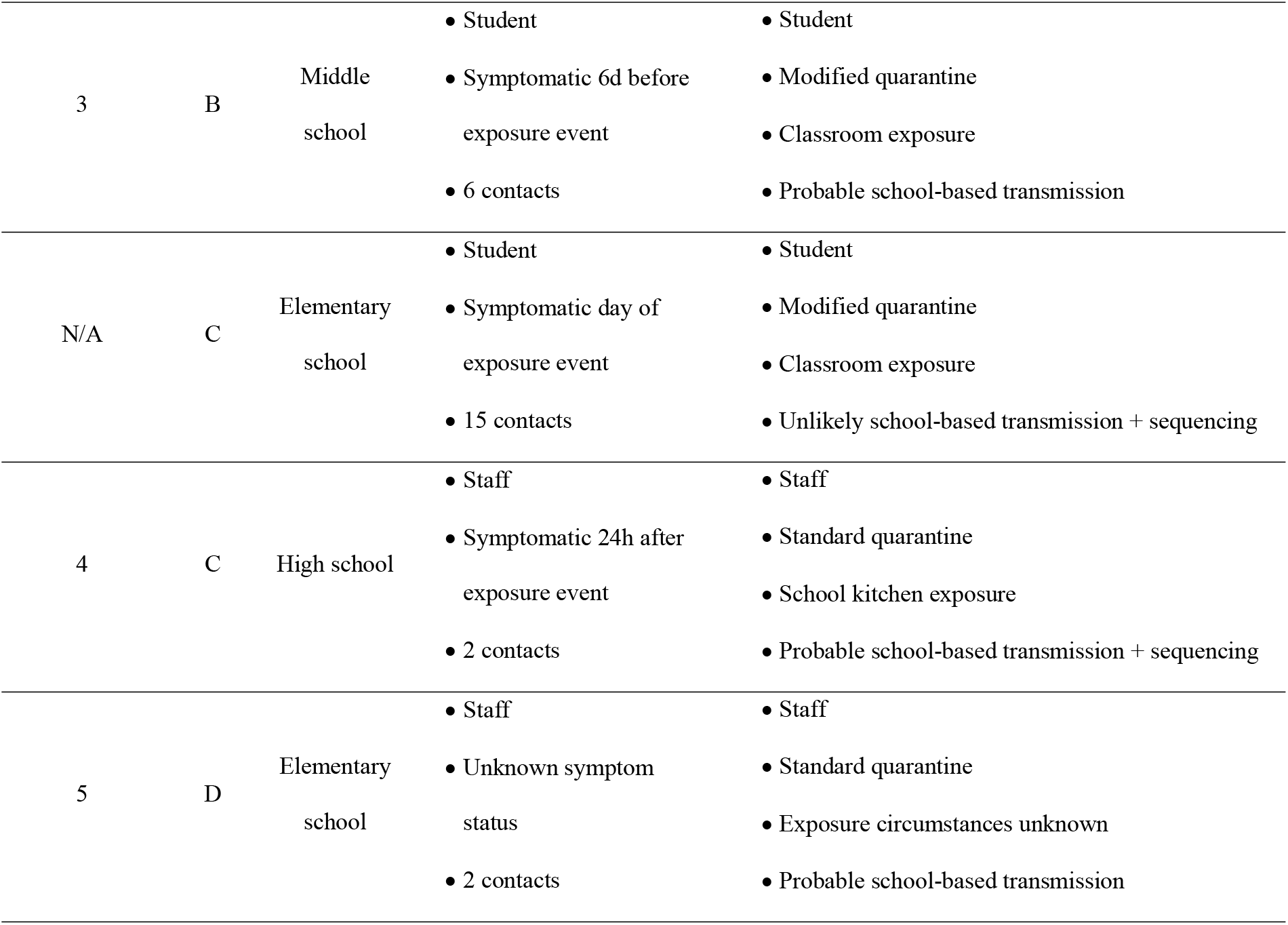
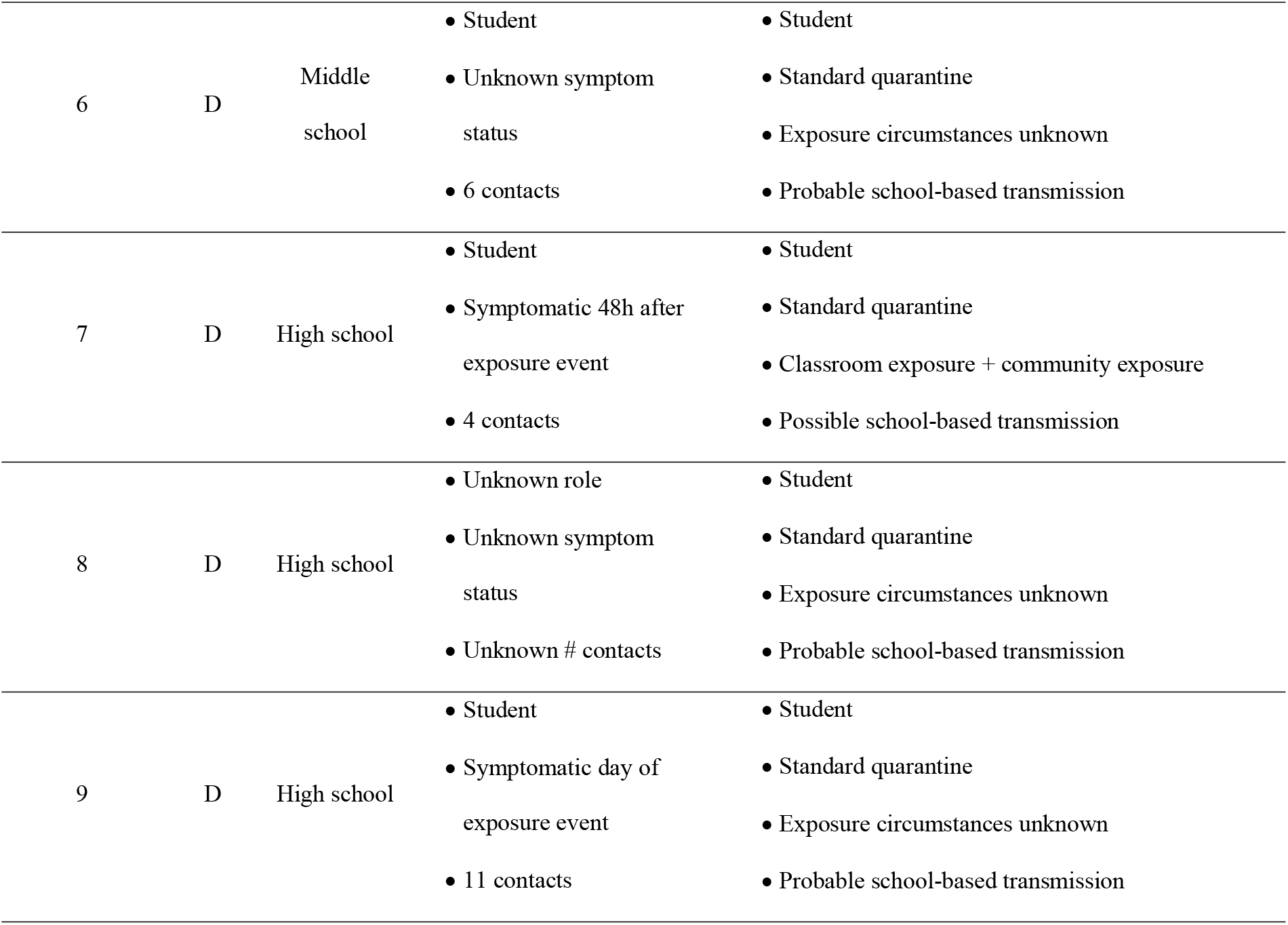
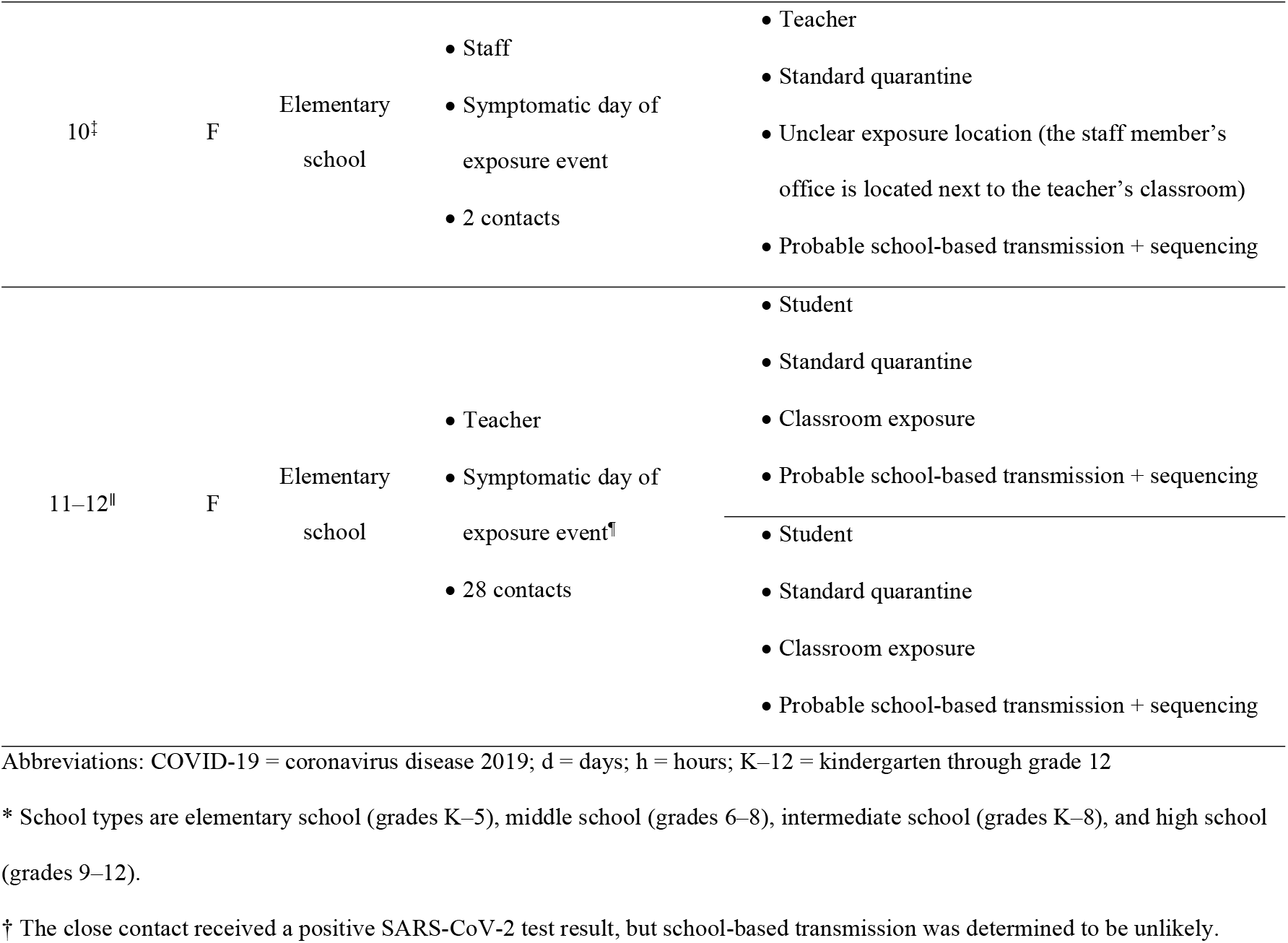
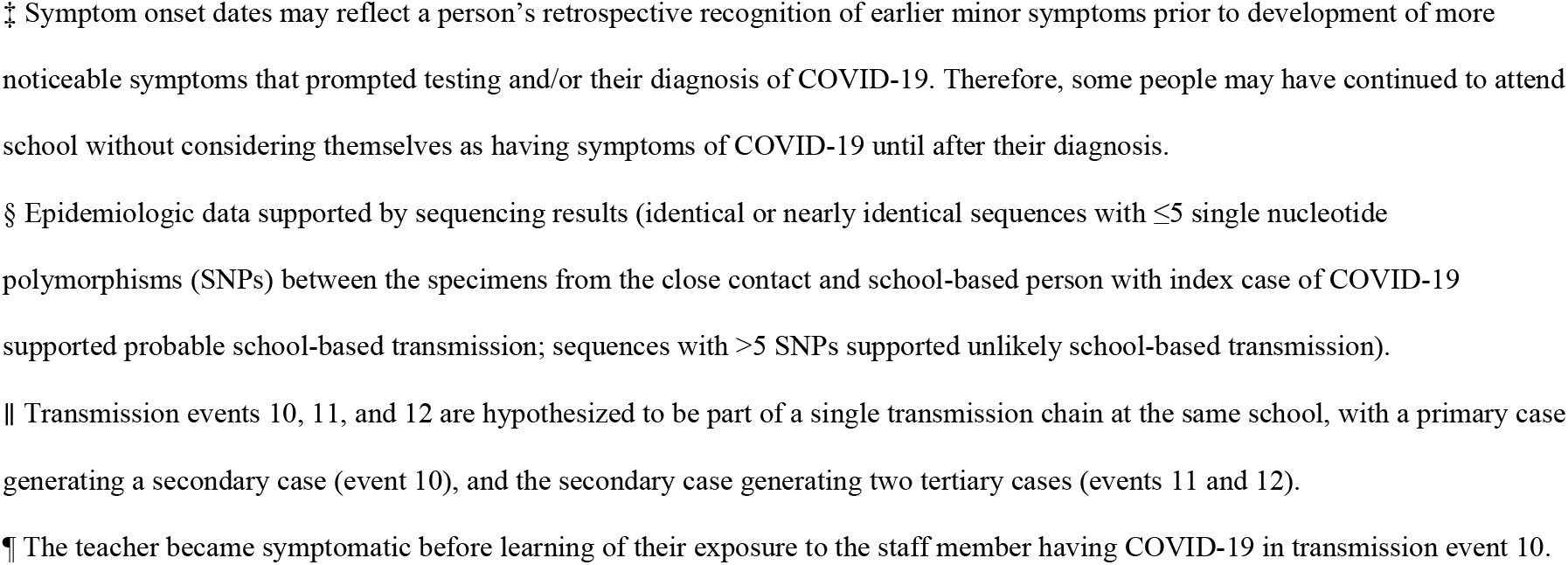
Characteristics of school-based persons with index cases of COVID-19 and close contacts who received a positive SARS-CoV-2 test result from K–12 schools, Greene and St. Louis Counties, Missouri, January 25–March 21, 2021.

Of 307 contacts we tested who reported no symptoms during the 14 days after their school-based exposure, 3 (1%) were positive. Therefore, among 1,140 contacts with no reported test results, a projected 11 additional asymptomatic infections may have occurred for a projected total of 23 (1%) school-based transmission events.

Unadjusted and adjusted relative risks of school-based SARS-CoV-2 infection among school contacts by schoolwide COVID-19 policy were statistically underpowered, primarily due to the low number of observed school-based infections and the homogeneity of prevention strategies implemented by schools (S2 Table).

In schools with a modified quarantine policy, 336 (49%) of 681 contacts were eligible for modified quarantine. Among 345 contacts who school officials deemed ineligible for modified quarantine, 29 were not eligible because they were not students. The primary reasons cited for ineligibility among 316 students include exposure during lunch (n=143; 45%), unmasked exposure (n=103; 33%), exposure during athletic activities (n=64; 20%), and prolonged direct contact (n=34; 11%); multiple reasons were cited for some students (Fig 1).

**Fig 1.**
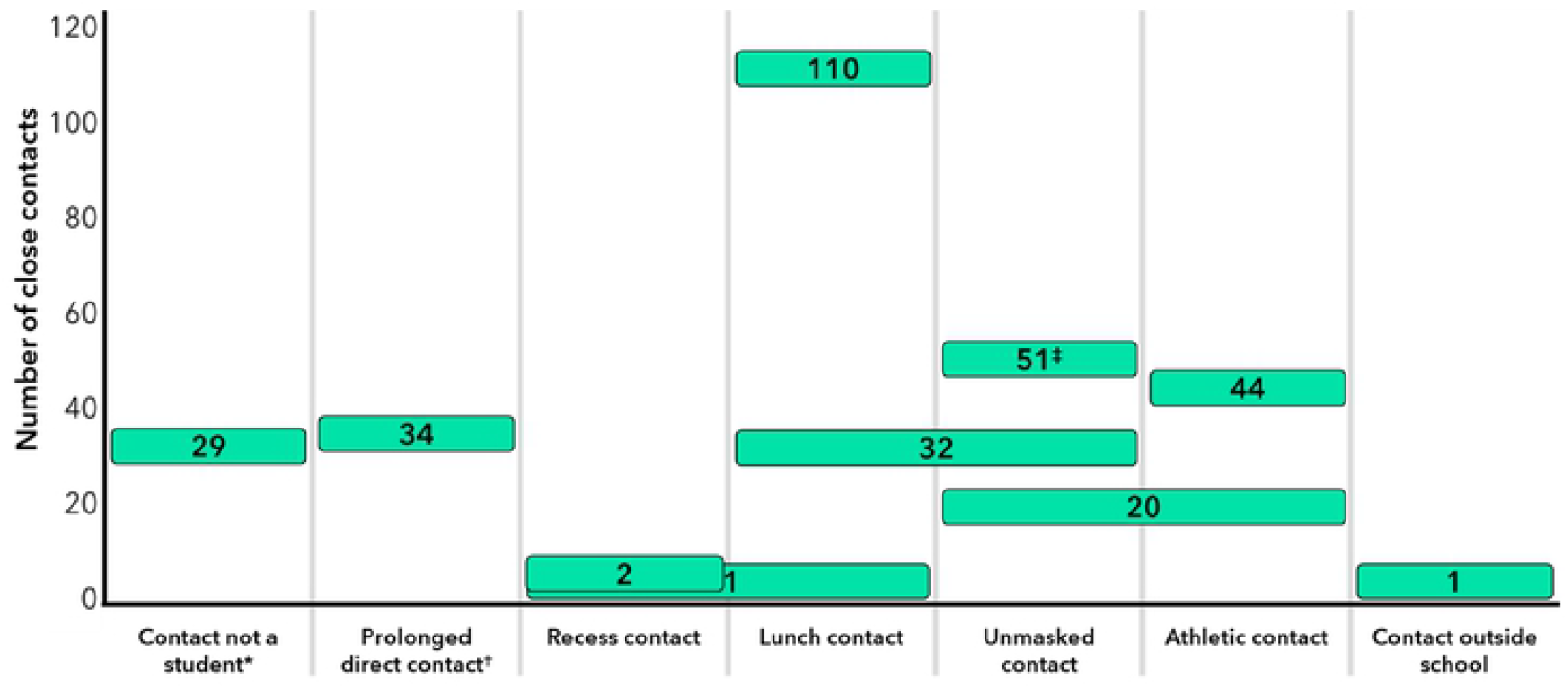
Primary reasons close contacts of persons with COVID-19 (n=345) were deemed ineligible for modified quarantine by K–12 school officials, Greene County, Missouri, January 25–March 21, 2021. Abbreviations: COVID-19 = coronavirus disease 2019; K–12 = kindergarten through grade 12 Caption: The number inside a rectangle corresponds to the number of close contacts who were deemed ineligible for a modified quarantine based on the category below the rectangle. Rectangles that overlap two reasons indicate both reasons were cited in the decision to deem the individual ineligible for a modified quarantine. *Under Greene County’s modified quarantine policy, only students aged ≤18 years were eligible for a modified quarantine. † Prolonged direct contact was defined as direct physical contact with the person having COVID-19 for ≥15 minutes. ‡ Includes 7 contacts who were also ineligible due to extracurricular activities and 2 contacts who were also ineligible due to contact outside of school.

A total of 66 (20%) students in a modified quarantine had available test results; three students with positive tests (5% of tested; 1% of all students in modified quarantine) were determined to have been infected through school-based transmission. All three students discontinued modified quarantine upon receiving a positive test result to begin isolation. None infected another person in the school environment; therefore, there was no observed onward transmission from students in modified quarantine. A projected additional three cases would be expected among the 270 students in modified quarantine without test results for a total of six (2%) transmission events among students in modified quarantine. A projected additional eight cases would be expected among the 835 students in standard quarantine without test results for a total of 14 (1%) transmission events among students in standard quarantine. The difference in frequency of transmission events between student contacts in modified versus standard quarantine was not different when comparing observed cases (*P*=0.41) or projected cases (*P*=0.50); thus, students selected for modified quarantine were no more likely to test positive or develop disease and pose a risk for onward school-based transmission than students in standard quarantine.

Using observed cases, the average crude incidence rate of school-based SARS-CoV-2 infections was 1.94 per 100,000 per week in schools that implemented a modified quarantine policy and 4.00 per 100,000 per week in schools following standard quarantine (*P*=0.24). The adjusted hazard rates of school-based SARS-CoV-2 infections were not different between schools that implemented a modified quarantine policy and schools that did not when using observed cases or total projected cases (for both, hazard ratio, HR=1.00; 95% confidence interval, CI: 0.97–1.03). The adjusted probability of school-based SARS-CoV-2 infections based on total projected cases reached a maximum of 0.83% (95% CI: 0.75–0.91%) by the end of the study (Fig 2).

**Fig 2.**
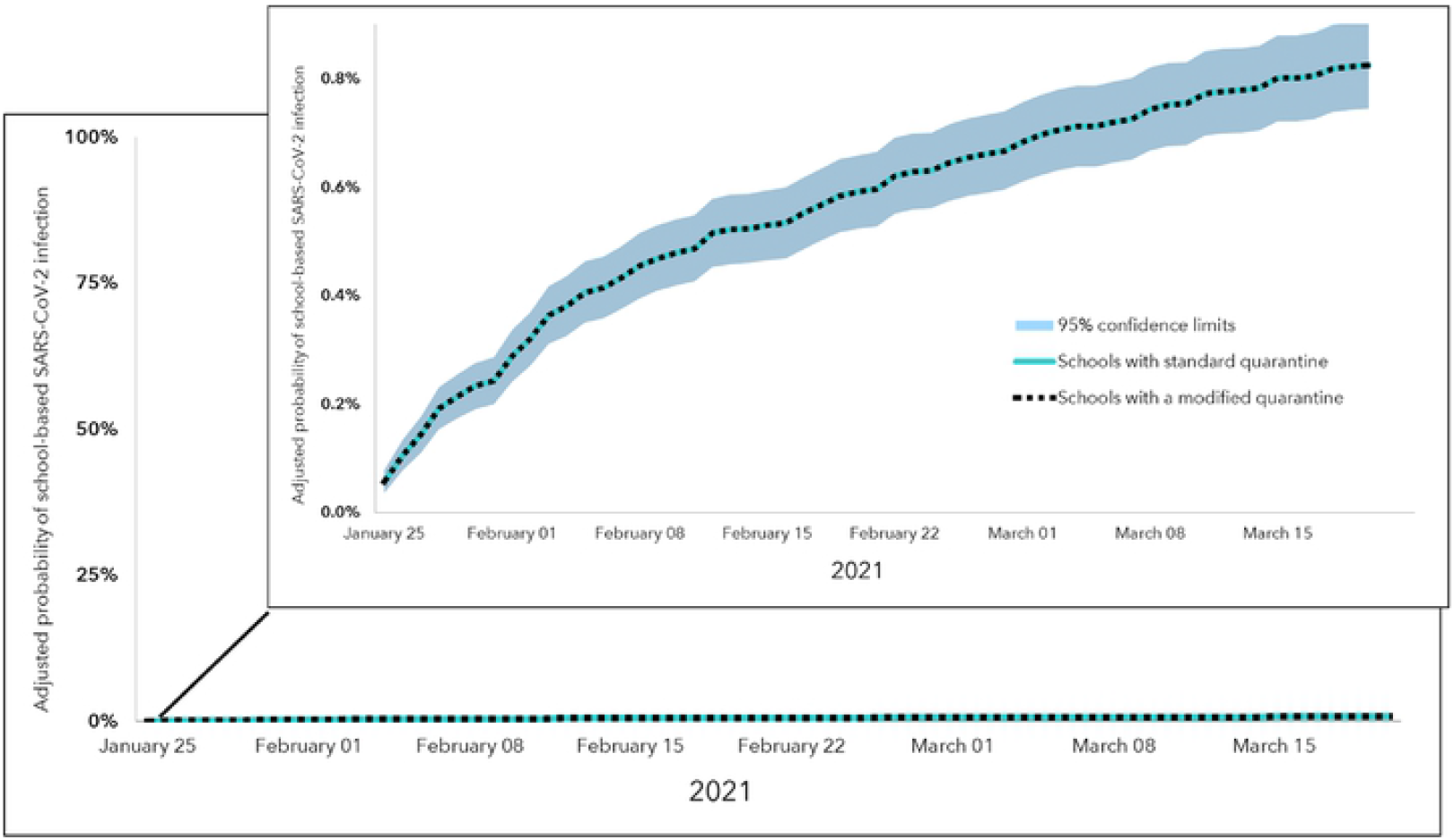
Adjusted probability of school-based SARS-CoV-2 infections based on total projected cases (n=23) in schools with and without a modified quarantine policy, Greene and St. Louis Counties, Missouri, January 25–March 21, 2021. Caption: Top-right inset shows zoomed-in view of adjusted probability curves over the study period. The adjusted* probability curve of school-based SARS-CoV-2 infection in schools with a standard quarantine policy is shown by the teal line; in schools with a modified quarantine policy, it is shown by the dotted black line. The adjusted probability curves and 95% confidence intervals were the same for schools with and without a modified quarantine policy. * The Cox regression model was adjusted for potential school-level confounding factors: quartiles of the percentage of students eligible to receive free or reduced-price lunch (as a proxy for school resources) and the school’s total number of cases that attended school or a school-related event during the study period.

## Discussion

From January 25 to March 21, 2021, school-based SARS-CoV-2 transmission was rare across 103 Missouri public schools, with a projected total of 23 transmission events. All schools implemented multiple COVID-19 prevention strategies, such as universal masking policies, spacing desks three feet apart, increasing ventilation in classrooms, and contact tracing with implementation of quarantine and isolation policies. These measures have been shown to limit school-based SARS-CoV-2 transmission in some specific settings (9, 12-15), but this report demonstrates this in urban, suburban, and rural public school districts; in elementary, middle, and high school grades; in schools that have implemented a modified quarantine and those that have not; during periods of moderate-to high-community incidence (21, 22); and for a combined estimated school population of >65,000 students, teachers, and staff. Since this investigation, there have been two waves of increased incidence of COVID-19 in the U.S. driven by the Delta and Omicron variants (23, 24). During future times of increased COVID-19 Community Levels(8), implementation of layered COVID-19 prevention strategies in schools will continue to play a pivotal role in preventing school-based SARS-CoV-2 transmission.

Modifications to student quarantine policies to allow students to continue in-person learning following a low-risk exposure to a person with COVID-19 at school did not result in onward SARS-CoV-2 transmission by generation of tertiary cases. These policies also did not result in increased schoolwide incidence relative to schools that did not implement a modified quarantine policy, even after adjusting for potential confounding factors. Student close contacts deemed eligible for modified quarantine were no more likely to receive a positive test result than their peers in standard quarantine and regained a combined estimated 2,664 days of in-person schooling (assuming each of the 333 students in modified quarantine who did not test positive would have missed eight school days if they had not been eligible for modified quarantine), enough school days for one student to attend kindergarten through their high school graduation. Other school districts with multiple prevention strategies in place have eliminated quarantine when masks are reliably used among persons with COVID-19 and their contacts without observing untoward effects (13). Beyond the educational benefits of restoring in-person learning, consideration of modified quarantine policies should factor in the psychosocial impacts on students and families.

There are several limitations to this report. First, school contact tracing may have not identified all persons exposed to someone with COVID-19 in a school setting. Second, we did not test all identified contacts for SARS-CoV-2 following exposure, and therefore the observed number and incidence of school-based SARS-CoV-2 infections is possibly an underestimation. However, positive test results outside of the enhanced investigation were reported to school officials and we projected the number of asymptomatic infections among those without test results to minimize underestimation. Third, due to low variability in school-level prevention strategies and the low number of identified school-based transmission events, analyses of the effect of specific prevention strategies on SARS-CoV-2 transmission were underpowered. Fourth, sequencing data and interview data were not available for all identified school index case-positive contact pairs, and in these instances, all persons who received a positive test result during the 14-day window were presumed to have been infected in school. For the six contacts with a positive test result who had paired sequencing data, it was determined that two (33%) were not infected from their school-based index case; it is possible that if sequencing data were available, school-based transmission may have been ruled out for some of the other contacts with a positive test result.

### Public Health Implications

In this two-month investigation of SARS-CoV-2 transmission in 103 schools implementing layered COVID-19 prevention strategies and modifications to quarantine policies, school-based SARS-CoV-2 transmission was rare and schools that implemented a modified quarantine policy did not have greater incidence of school-based SARS-CoV-2 infections. Given these findings and the benefits of restoring in-person learning for students, schools implementing multiple COVID-19 prevention strategies might consider adopting a similar policy during times of increased community incidence. This should be done in conjunction with reinforcement of public health messaging to promote quarantine adherence outside of school. K–12 schools implementing universal face mask policies, promoting physical distancing, and increasing ventilation in classrooms continue to experience low rates of school-based SARS-CoV-2 transmission. As vaccination become available for the majority of the school-aged population since the completion of this investigation, additional consideration and adaptation to modified quarantine policies could be considered. Modifications to student quarantine policies may reduce disruptions to in-person education while maintaining a safe environment for the students and staff.

## Data Availability

Data cannot be shared publicly because of participation of students under the age of 18. Data are available from the Washington University in St. Louis Study PI (contact) for researchers who meet the criteria for access to confidential data.

## Supporting information

**S1 Table. Characteristics of 103 public K–12 schools participating in SARS-CoV-2 transmission investigation, Greene and St. Louis Counties, Missouri, January 25–March 21, 2021**

Abbreviations: K–12 = kindergarten through grade 12; NR = not reported

Note: Data are from surveys completed by school and district officials unless otherwise noted.

* Data from: https://dese.mo.gov/school-data

† Includes participating schools only.

‡ Race and ethnicity categories at the school level differed from those collected from individuals as part of the investigation.

**S2 Table. Unadjusted and adjusted relative risks of school-based SARS-CoV-2 infection among school close contacts by schoolwide COVID-19 policy in K–12 schools, Greene and St. Louis Counties, Missouri, January 25–March 21, 2021**.

Abbreviations: COVID-19 = coronavirus disease 2019; K–12 = kindergarten through grade 12; RR = relative risk; CI = confidence interval

Note: All regression models accounted for cluster-correlated observations at the school level using generalized estimating equations (GEE) with an independent correlation matrix.

* egression models were adjusted for school role (student versus staff).

† Surveyed ventilation strategies included opening doors when possible, opening windows when possible, using fans to circulate air, and updating heating, ventilation, and air conditioning (HVAC) systems specifically to prevent COVID-19.

**S1 Fig. Schoolwide COVID-19 policies reported in 100 K–12 schools, Greene and St. Louis Counties, Missouri, January 25–March 21, 2021**.

Abbreviations: COVID-19 = coronavirus disease 2019; HVAC = heating, ventilation, and air conditioning; K–12 = kindergarten through grade 12

* School buildings also include the areas listed in the subsequent two categories: hallways, stairways, gymnasiums, cafeterias, and other special use rooms.

## Acknowledgments

All the students, families, educators, nurses, administrators, and staff members from participating schools and school districts in Greene and St. Louis Counties, Missouri; Dr. James Blaine; Pam A. Miller; Dr. Matt Pearce; Dr. Shawn Randles; CDC COVID-19 Response Team (Jennifer Frazier, Elizabeth Haller, Laura Hughes-Baker, Catherine N. Rasberry, Hailey Reid); Jordan Valley Community Health Center (Shelley Hall, Shannel Johnson, Mandy Jones, Jennifer Logan, Dr. Matthew Stinson, Amber Tenorio); Mercy Hospital Springfield and St. Louis; Ozarks Technical Community College (Katelyn Antrim, Dr. Daniela Brink, Alane Cordray, Katherine Ebersole, Chanté Whittle); Saint Louis University (Mary Beal, Adrienne Beckett-Ansa, Janaki Bhave, Allie Bodin, Kajal Dholakia, Andrea Hoppert, Rachel Leimkuehler, Elena Dalleo Locascio, Ruband Mahmood, Rachel L. Mazzara, Margaret O’Brien, Komal Patel, Lydia Thomas, Audrey Yao); Springfield-Greene County Health Department (Sean Barnhill, Jordan Coiner, Brad Stulce, Kathryn Wall); University of Missouri School of Medicine (Hosea Covington, Evan Garrad, Dr. David Haustein, Tricia Haynes, Spencer Blake Price, Wyatt Whitman); 4ES Corporation (Dr. Bre Peeler); and Washington University in St. Louis (Lori Barganier, Caleb Gentry, Ian T. Lackey, Savanah Low, Dr. Suong T. Nguyen, Jenna Rideout, Tejas Sekhar, Cindy Terrill, Cole Tipton).

## COVID-19 Response Fieldwork and Laboratory Teams

Thu-ha Dinh, M.D., Catherine V. Donovan, Ph.D., Victoria Foltz, B.S., Jessica L. Halpin, M.S., Sooji Lee, M.S.P.H., Michelle O’Hegarty, Ph.D., Jonathan Steinberg, M.P.H., Elaine Stevens-Emilien, M.S., Shaniece C. Theodore, Ph.D., Caitlin M. Worrell, M.P.H. (COVID-19 School Fieldwork Team, CDC); Yan Li, M.Sc., Ying Tao, Ph.D., Anna Uehara, Ph.D., Jing Zhang, Ph.D. (COVID-19 Surge Laboratory Group, CDC); Sarah Greene, M.D., Ph.D., Jaimee Hall, D.O., Alex S. Plattner, M.D. (COVID-19 School Fieldwork Team, Washington University in St. Louis).

## Missouri School District Data and Coordination Group

Tammy Fitzpatrick, B.S.N., R.N., Mandy Williams, R.N., Logan-Rogersville School District, Rogersville, MO; Amanda Fields, B.S.N., R.N., Tim Pecoraro, Ed.D., Pattonville School District, St. Ann, MO; Natalie Botkin, B.S.N., R.N., Republic School District, Republic, MO; Glenn Hancock, Ed.S., Amy Wehr, B.S.N., R.N., Rockwood School District, Eureka, MO; Jean Grabeel, M.Ed., R.N., Lee Ann Neill, M.S.N., R.N., Springfield Public Schools, Springfield, MO; Kashina Bell, Ed.D., Sharonica Hardin-Bartley, Ph.D., The School District of University City, University City, MO.

